# Differentially Altered Metabolic Pathways in the Amygdala of Subjects with Schizophrenia, Bipolar Disorder and Major Depressive Disorder

**DOI:** 10.1101/2024.04.17.24305854

**Authors:** Xiaolu Zhang, Jake Valeri, Mahmoud A. Eladawi, Barbara Gisabella, Michael R. Garrett, Eric J Vallender, Robert McCullumsmith, Harry Pantazopoulos, Sinead M. O’Donovan

**Author notes:** **Address for correspondence:** Sinead O’Donovan, PhD, Department of Neuroscience, University of Toledo Medical Center, 183 Block Health Science Building, 3000 Transverse Dr., Toledo, OH 43614, Harry Pantazopoulos, PhD, Department of Psychiatry and Human Behavior, University of Mississippi School of Medicine, TRC 407, 2500 North State St, Jackson, MS 39216, USA.

## Abstract

**Background and hypothesis:** A growing number of studies implicate a key role for metabolic processes in psychiatric disorders. Recent studies suggest that ketogenic diet may be therapeutically effective for subgroups of people with schizophrenia (SCZ), bipolar disorder (BPD) and possibly major depressive disorder (MDD). Despite this promise, there is currently limited information regarding brain energy metabolism pathways across these disorders, limiting our understanding of how brain metabolic pathways are altered and who may benefit from ketogenic diets. We conducted gene expression profiling on the amygdala, a key region involved in in the regulation of mood and appetitive behaviors, to test the hypothesis that amygdala metabolic pathways are differentially altered between these disorders.

**Study Design:** We used a cohort of subjects diagnosed with SCZ, BPD or MDD, and non-psychiatrically ill control subjects (n=15/group), together with our bioinformatic 3-pod analysis consisting of full transcriptome pathway analysis, targeted pathway analysis, leading-edge gene analysis and iLINCS perturbagen analysis.

**Study Results:** We identified differential expression of metabolic pathways in each disorder. Subjects with SCZ displayed downregulation of mitochondrial respiration and nucleotide metabolism pathways. In comparison, we observed upregulation of mitochondrial respiration pathways in subjects with MDD, while subjects with BPD displayed enrichment of pathways involved in carbohydrate metabolism. Several pathways associated with brain metabolism including immune system processes and calcium ion transport were also differentially altered between diagnosis groups.

**Conclusion:** Our findings suggest metabolic pathways are differentially altered in the amygdala in these disorders, which may impact approaches for therapeutic strategies.

## INTRODUCTION

A growing number of studies highlight a high degree of comorbidity of metabolic disorders in people with schizophrenia (SCZ), bipolar disorder (BPD) or major depressive disorder (MDD), including cardiovascular disease and diabetes ^1–6^. Comorbid metabolic syndrome negatively impacts treatment strategies and life expectancy in people suffering from these psychiatric disorders ^7^. Furthermore, the prevalence of metabolic syndrome has been increasing in the U.S., with approximately 34.7% of the population meeting diagnostic criteria ^8^. Recent studies suggest that brain metabolic dysfunction is associated with specific psychiatric symptoms, including cognitive dysfunction, mania, depression and psychosis, as well as with effects of psychotropic medications ^9–14^. For example, metabolic dysregulation is associated with cognitive dysfunction in people with mood disorders ^14^, and alterations in molecular signaling pathways in glucose regulation have been reported recently in antipsychotic naïve patients with psychosis ^15^. Furthermore, a meta-analysis of genetic factors from GWAS studies and candidate gene studies identified twenty-four genetic factors that are shared between subjects with mood disorders and subjects with metabolic disorders ^16^.

A number of recent clinical and preclinical studies have highlighted a key role of systemic metabolic dysfunction in the pathophysiology and treatment of a range of psychiatric disorders, including SCZ, BPD and MDD ^17–20^. Impaired glucose metabolism and mitochondrial dysfunction are proposed as core features of these disorders. The ketogenic diet, originally proposed as a treatment for epilepsy ^21^, is emerging as an effective treatment for psychiatric disorders ^20, 22–25^ and may address dysfunction in glucose metabolism and mitochondrial processes ^17–20^.

Several studies provide evidence for brain metabolic abnormalities in subjects with mood disorders and/or psychosis. Recent gene expression profiling studies from prefrontal cortex samples identified alterations in pathways involved in mitochondrial function and oxidative phosphorylation in subjects with SCZ ^26^. Electron microscopy analysis of postmortem prefrontal cortex samples identified altered mitochondrial architecture in subjects with BPD ^27^. Furthermore, bioinformatic analysis on transcriptomic data from peripheral tissues identified altered pathways in glucose signaling in subjects with psychosis ^15^.

Despite the support for brain metabolic dysfunction in psychiatric disorders, evidence comparing metabolic signaling pathways in brain areas critically involved in mood and reward processes in subjects with SCZ, BPD, or MDD is limited. The amygdala is a key region involved in the regulation of mood, anxiety, fear, and reward (appetitive) processes. Imaging studies reported increased amygdala activity across each of these disorders ^28–30^. For example, increased amygdala activity was reported in unmedicated subjects with paranoid SCZ ^28^ and in subjects with BPD including mixed-mania and rapid cycling BPD ^29^. Similarly, increases in amygdala glucose metabolism in subjects with depression or BPD correlated with plasma cortisol levels ^30^.

Several studies indicate that the amygdala is a brain region where neuroanatomical and molecular pathology diverges in psychiatric disorders. Differences in resting state functional connectivity between the amygdala and prefrontal cortex are reported in subjects with SCZ and subjects with BPD ^31^. Amygdala volume is decreased in children and adults with BPD ^32^ and total neuron number is also decreased in BPD but not SCZ subjects ^33^. In comparison, amygdala volume is increased in subjects with MDD ^34^. In addition, decreased perineuronal nets and marked increases of chondroitin sulphate proteoglycan-expressing glial cells were reported in the amygdala of subjects with SCZ but not in subjects with BPD ^35^.

Improving our understanding of how brain metabolic processes are affected in people with SCZ, BPD or MDD can provide insight into the use of current as well as new treatment strategies for people with and without comorbid metabolic syndrome. There is currently a lack of studies comparing amygdala gene expression profiles across these three disorders. We used RNAseq profiling and our 3-pod bioinformatic analysis on a well-characterized cohort from the Stanley Neuropathology Consortium to determine whether metabolic signaling pathways are differentially altered in the amygdala of subjects with SCZ, BPD or MDD.

## METHODS

### Subjects

Fresh frozen coronal sections containing the amygdala were obtained from the Stanley Neuropathology Consortium. This cohort consists of 15 subjects with SCZ, 15 subjects with BPD, 15 subjects with MDD and 15 control subjects (**Table 1**). Details regarding this cohort including demographic and patient information is available at the <u>Stanley Neuropathology Consortium</u> <u>Integrative Database</u>. Ethical approval for the Stanley Brain Collection was obtained through the Uniformed Services University of the Health Sciences, Bethesda, MD.

**Table 1:** Summary table of subjects and demographic information. C: Caucasian, A: Asian, A.A.: African American. R: right, L: left. M: male, F: female. Information for all demographic and patient information used for our analysis is available at the <u>Stanley Neuropathology Consortium Integrative Database</u>.

|  | Schizophrenia | Bipolar Disorder | Major Depressive Disorder | Control |
| --- | --- | --- | --- | --- |
| **Age (years)** | 44.2 (25-62) | 42.3 (25-61) | 46.5 (30-65) | 48.1 (29-68) |
| **Sex** | 9 M, 6 F | 9 M, 6 F | 9 M, 6 F | 9 M, 6 F |
| **Race** | 12 C, 3 A | 14 C, 1 AA | 15 C | 14 C, 1 AA |
| **PMI (hrs)** | 33.7 (12-61) | 32.5 (13-62) | 27.5 (7-47) | 23.7 (8-42) |
| **pH** | 6.1 (5.8-6.6) | 6.2 (5.8-6.5) | 6.2 (5.6-6.5) | 6.3 (5.8-6.6) |
| **Hemisphere** | 6 R, 9 L | 8 R, 7 L | 6 R, 9 L | 7 R, 8 L |
| **Suicide** | 11 No, 4 Yes | 6 No, 9 Yes | 8 No, 7 Yes | 15 No, 0 Yes |

### RNAseq & alignment

RNA isolation, library preparation, and next generation sequencing was performed by the Molecular and Genomics Core Facility at the University of Mississippi Medical Center, as described previously ^36^. Total RNA was isolated from tissue samples using the Invitrogen PureLink RNA Mini kit with Trizol (Life Technologies; Carlsbad, CA, USA) following manufacturer protocol. Quality control of total RNA was assessed using the Qiagen QIAxcel Advanced System for quality and Qubit Fluorometer for concentration measures. The RQI was 6.6 ± 2.1 (mean ± SD). Libraries were prepared using the TruSeq Stranded Total RNA LT Sample Prep Kit from Illumina (San Diego, CA, USA) per manufacturer’s protocol using up to 1 ug of RNA per sample. Libraries were index-tagged, pooled for multiplexing, and sequencing was performed on the Illumina NextSeq 500 platform using a paired-end read (2 x75 bp) protocol with the Illumina 150 cycle High-Output reagent kit. Reads were aligned to the NCBI GRCh38Decoy Refseq genome with the basespace application RNA-Seq Alignment (Version: 2.0.1 [workflow version 3.19.1.12+master]) that conducted both splice aware genome alignment with STAR alignment (version 2.6.1a, ^37^) and transcriptome quantification with Salmon (version 0.11.2) ^38^.

### Differential gene expression analysis

To better understand the drivers of expression variation in our study, variance partition analysis was conducted using variancePartition R package ^39^. This analysis allowed us to quantify the variation in each expression trait that could be attributed to differences in each covariate. Each subject in the study was associated with sixteen covariates, including sex, age, duration of illness, age at illness onset, cause of death suicide, psychosis, lifetime antipsychotics (fluphenazine equivalent), smoking history, substance abuse severity, ethanol severity, diagnosis, zeitgeber time (ZT), brain hemisphere, tissue pH, brain weight, and PMI. The top two covariates with the greatest median were accounted for in the subsequent generalized linear modeling fitting for each comparison. Transcriptome-wide gene counts were subject to differential gene expression analysis using DEseq2 R package ^40^ with recommended default settings. Genes where there are less than 50% samples with normalized counts greater than or equal to 1 were filtered out. Unless otherwise specified, significantly differentially expressed genes are defined as those with a p-value less than 0.05.

### Pod 1: Full gene set pathway analysis

Gene Set Enrichment Analysis (GSEA) with full set of genes was performed using fgsea R package (version 1.16.0) against human enrichment map gene sets downloaded from baderlab.org/ EM_Genesets/. As a gene ranking metric, sign(logFC) * (−log10(p-value)) or “stat” obtained from DESeq2 output were used. The GSEA method is described in detail ^41^. Briefly, GSEA first ranks genes based on differential expression. Then an enrichment score statistic is generated, which reflects the degree of overrepresentation of genes in a gene set at the top or bottom of the entire list of ranked genes. Unless otherwise specified, significantly altered pathways (gene sets) are defined as those with a p-value less than 0.05. This analysis also identified leading−edge (LE) genes which are the core subset of genes in a gene set that account for the enrichment signal ^41^. We analyze the overlap between multiple leading-edge subsets.

### Pod 2: Targeted pathway analysis

Targeted pathway analysis with disease gene sets composed of the top and bottom 10% genes (greatest absolute log2FC) was performed using enrichR R package (version 3.0). Gene ontology (GO) databases GO biological process, GO cellular component, and GO molecular function were used in analysis. EnrichR generates a combined score to identify pathways that are significantly up or downregulated from a given list of DEGs ^42^.

### Pod 3: Identification of perturbagens altering gene expression

The Library of Integrated Network-Based Cellular Signatures (LINCS) (http://www.ilincs.org/ilincs/) is a National Institute of Health initiative that aims to create a comprehensive network of molecular reactions in response to environmental and internal stressors ^43^. The LINCS project uses the L1000 assay, a gene expression array of 978 “hub” genes, to generate gene signatures. Approximately 82% of the information content of the transcriptome is accounted for in the genes represented in the L1000 assay ^44^. The LINCS database contains hundreds of thousands of gene signatures, including gene signatures generated in human cell lines treated with chemical perturbagens (drugs). The log2FC and p-value for the L1000 genes were extracted from DEG analysis (disease signatures) and submitted as input to inquire a list of chemical perturbagens (drug signatures). The reported score is the Pearson correlation coefficient between the disease signatures (SCZ v CTL, BPD v CTL, MDD v CTL) and the precomputed iLINCS drug signatures. The chemical perturbagens with discordance scores < −0.321 and concordance scores > 0.321 were retained. Chemical perturbagens were clustered by mechanism of action (MOA) categories inquired from L1000 FWD ^45^, DrugBank database ^46^, and the Broad Institute. The R script for the 3-pod DEG workflow incorporating GSEA, targeted pathway analysis and iLINCS analysis is available at https://zenodo.org/badge/latestdoi/642681935.

### Psychotropic medication gene overlap analysis

To determine whether the DEG changes observed in this study were affected by psychotropic medications, a hypergeometric test was performed using GeneOverlap R package (version 3.17) (https://github.com/shenlab-sinai/GeneOverlap) to compare the neuropsychiatric disorder differential expression profiles (filtered by FDR<0.01, FDR<0.05, and p-value <0.05) to transcriptomic datasets (filtered by p-value < 0.05) obtained from the Stanley Online Genomics resource (https://www.stanleygenomics.org/) for SCZ subjects who were on and off antipsychotic medication and BPD subjects on and off mood stabilizers at time of death. The Jaccard Index, which assesses the similarity between two sets of genes and p-value of the statistic test, is reported as previously described ^47^.

### Cell type deconvolution analysis

This study leveraged single-cell expression profiles obtained from snRNA-seq data of human amygdala collected from eight postmortem brain donors by the Lieber Institute for Brain Development ^48^. The dataset includes 19 clusters representing glial, stromal, immune cell populations, and neuronal classes. The raw counts and annotated single-cell clusters were obtained as SingleCellExperiment objects from (https://github.com/LieberInstitute/10xPilot_snRNAseq-human). The snRNA-seq raw counts data underwent filtering to remove genes with zero expression across all cells. Subsequently, normalization was performed using the Trimmed Mean of M-values with singleton pairing (TMMwsp) method from the edgeR package ^49^, suitable for data with a high proportion of zeros. Cellular deconvolution was performed using CIBERSORTx ^50^, employing support vector regression (SVR) for cellular proportions estimation. To create a signature matrix containing marker genes for each cell type, the *cibersortxfractions* docker container was used, with adjustments made for the microfluidics-based sequencing (10xGenomics) by setting the fraction parameter to 0 as recommended by ^50^. To enhance visualization, cell types with predominantly zero fractions across all samples were excluded. Excitatory and inhibitory neuron subcluster fractions were combined into their respective classes. Additionally, the proportions of each cell type across all groups were normalized to the mean of the control group. The significance of altered cell-type proportions among the different groups was assessed using the Wilcoxon Sum test.

## RESULTS

### Amygdala transcriptional signature in SCZ, BPD and MDD

Differential gene expression analysis identified 2,890 (SCZ), 3,795 (BPD) and 3,016 (MDD) nominally significant differentially expressed gene (DEGs) with p-value < 0.05 (volcano plot **Figure S1, Table S1**). DEGs included SERPINA3, COL1A1, and OXTR in SCZ, CRY1, PPP3CC and PDYN in MDD, ABCG2, SERPINA3, several L-type calcium genes including CACNB2 and CACNA1C, and several extracellular matrix genes including NCAN PTPRZ1, VCAN, IL33, and several clock genes including PER3, PER1, PER2, and NPAS4 in BPD.

Variance partitioning analysis quantified the variation in each expression trait that could be attributed to differences in each covariate (% variance explained) (**Figure S2**). The top two covariates with the greatest median of variation for each disease were duration of illness (7.4%) and age at onset of illness (4.2%) for BPD; age at onset of illness (5.4%) and pH (4.1%) for MDD and age at onset of illness (11.2%) and duration of illness (4.1%) for SCZ comparisons.

### Pathway analysis

Pathway analysis (**Figure 1, Table S2**) conducted using GSEA takes advantage of information on continuous expression changes from all transcribed genes to determine the biological processes (gene sets) that are statistically significantly different between the disorder and control groups ^41^. Enrichment of pathways involved in “metabolic processes” (see pop-out cluster in **Figure 1**), were identified in all disorders. “Nucleotide metabolism” was downregulated in SCZ, with few upregulated pathways enriched in MDD. “Carbohydrate metabolism-related processes” were predominantly enriched in BPD. “Mitochondrial respiration/energy metabolism” related pathways were downregulated in SCZ, upregulated in MDD, but not significantly enriched in BPD. In line with previous reports, these results suggest dysregulation of bioenergetic processes in psychiatric disorders. However, they also indicate unique dysfunction in different energy metabolism pathways in the amygdala in these disorders. Similarly, “immune system processes” are enriched in all disorders but pathways are primarily upregulated in BPD and downregulated in SCZ (**Figure 2**). Fewer immune-related pathways are enriched in MDD in the amygdala compared to BPD and SCZ.

**Figure 1.**
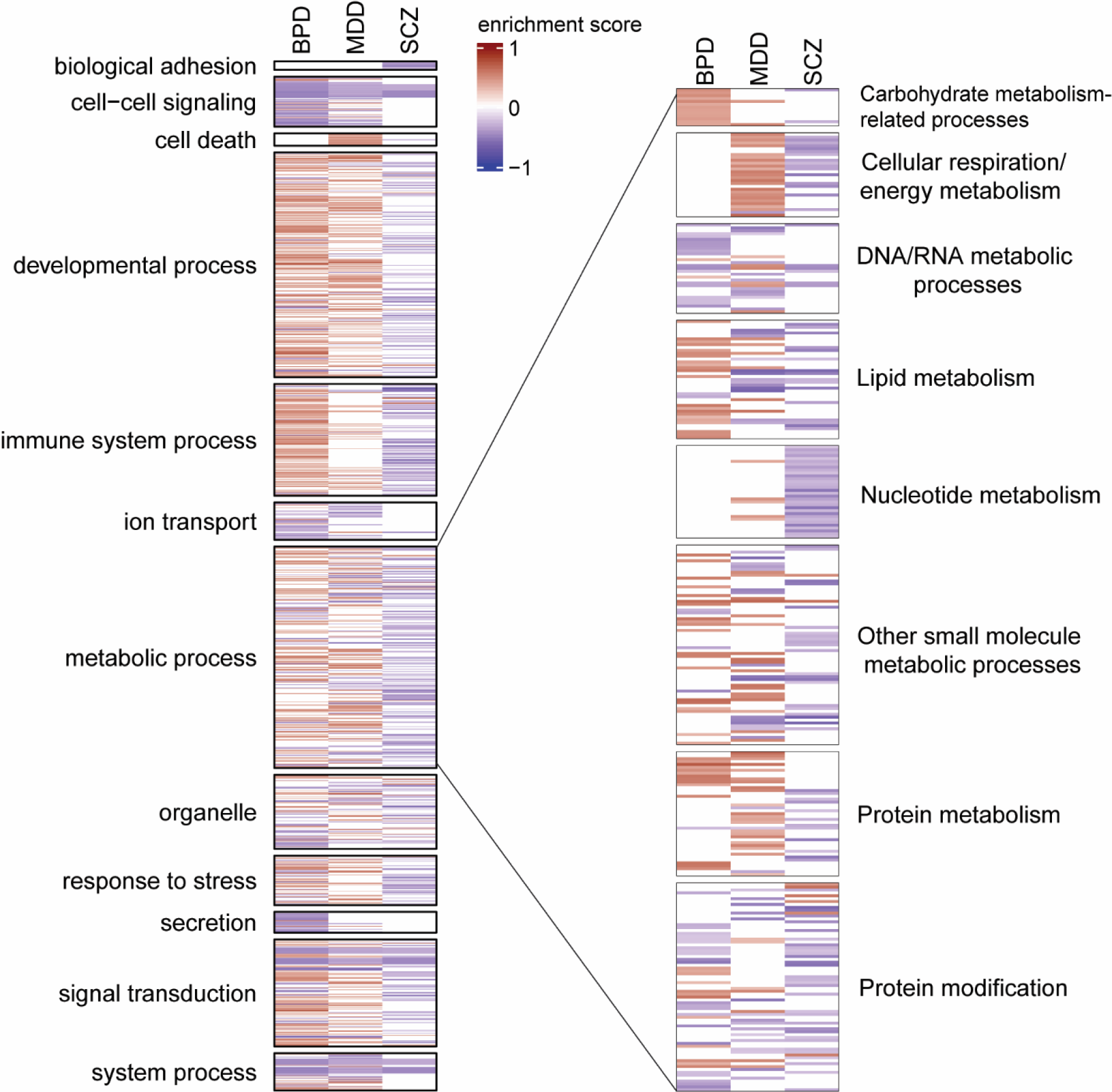
Pathway analysis of DEGs in amygdala in three psychiatric disorders. Enrichment of clustered biological pathways following gene set enrichment analysis (GSEA) of BPD, MDD and SCZ comparisons. The color-intensity (blue to red) is proportional to the enrichment score (ES). The enrichment score represents the degree to which the genes in the set are over-represented at either the top or bottom of the list. The “metabolic processes” cluster is expanded. All pathways indicated in heatmap are significantly (p<0.05) enriched. BPD bipolar disorder, MDD major depressive disorder, SCZ schizophrenia.

**Figure 2.**
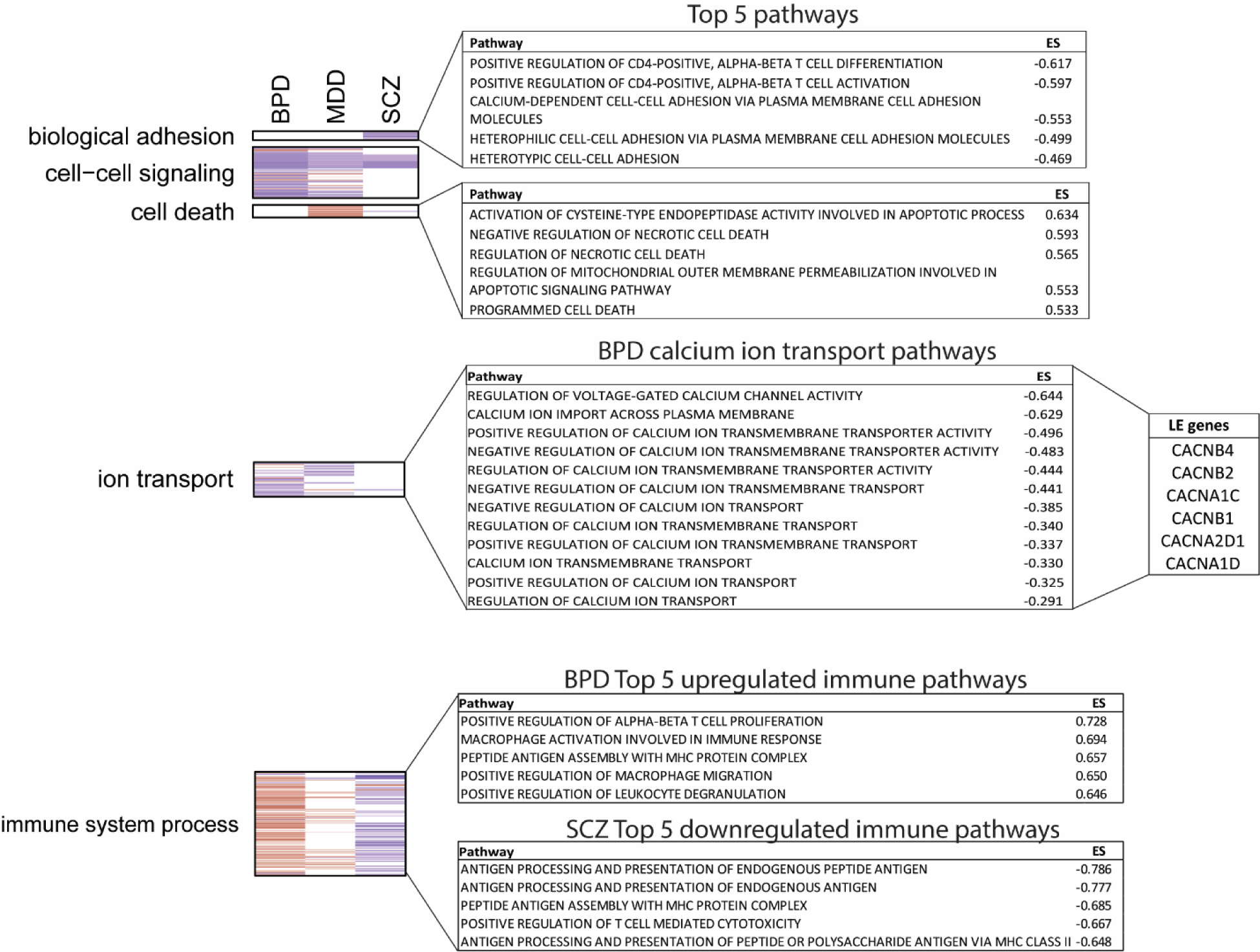
Pathway analysis. Heatmap indicating significantly (p<0.05) enriched pathway clusters that are predominately or uniquely enriched in a single psychiatric disorder, and table of corresponding top 5 pathways in each cluster. “Biological adhesion” and “cell-death” related pathways are enriched in SCZ and MDD respectively “Immune system process” pathways are upregulated in SCZ and downregulated in BPD. The top 5 pathways in each cluster are listed by enrichment score (ES). The top BPD “Ion transport” pathways and leading edge (LE) genes are also shown. BPD bipolar disorder, MDD major depressive disorder, SCZ schizophrenia.

Other pathway clusters contained pathways that were enriched across psychiatric disorders, however, pathways associated with “biological adhesion” and “cell death” were uniquely or predominantly downregulated in SCZ and upregulated in MDD, respectively (**Figure 1**). The top 5 pathways in these clusters, based on enrichment score (ES), are indicated in **Figure 2**. Furthermore, “calcium ion transport” pathways were predominantly downregulated in subjects with BPD. Several genes encoding for L-type calcium channels were identified as leading-edge genes (**Figure 2)** in these pathways, including CACNA1C, implicated in GWAS studies of BPD ^51^, and CACNB2, which has genetic associations with BPD ^52^ and with cardiovascular disease ^53, 54^. The top “calcium ion transport” pathways identified in BPD are listed in **Figure 2**.

### Leading-edge gene analysis

Leading-edge gene analysis identified the genes that are most influential for enrichment of significant pathways (**Table S3**). Leading-edge genes are identified based on the frequency with which they are identified in biological pathways; their expression is not necessarily statistically significant in disease compared to control. The top 10 upregulated and downregulated leading-edge genes are identified for BPD (**Figure 3A**), MDD (**Figure 3B**) and SCZ (**Figure 3C**). Approximately 11% (upregulated) and 15% (downregulated) of the leading-edge genes are shared across all 3 psychiatric disorders (**Figure 3D**). The highest number of common leading-edge genes are shared between SCZ and MDD (approx. 16%). BPD has the greatest number (approx. 24%) of unique LE genes. The top 10 common leading-edge genes (**Figure 3E**) include genes involved in metabolic processes (GPER1 & PPP3CA), L-type voltage gated calcium signaling (CACNB4), mu opioid signaling (OPRM1) and GABAergic signaling (GABRB2).

**Figure 3.**
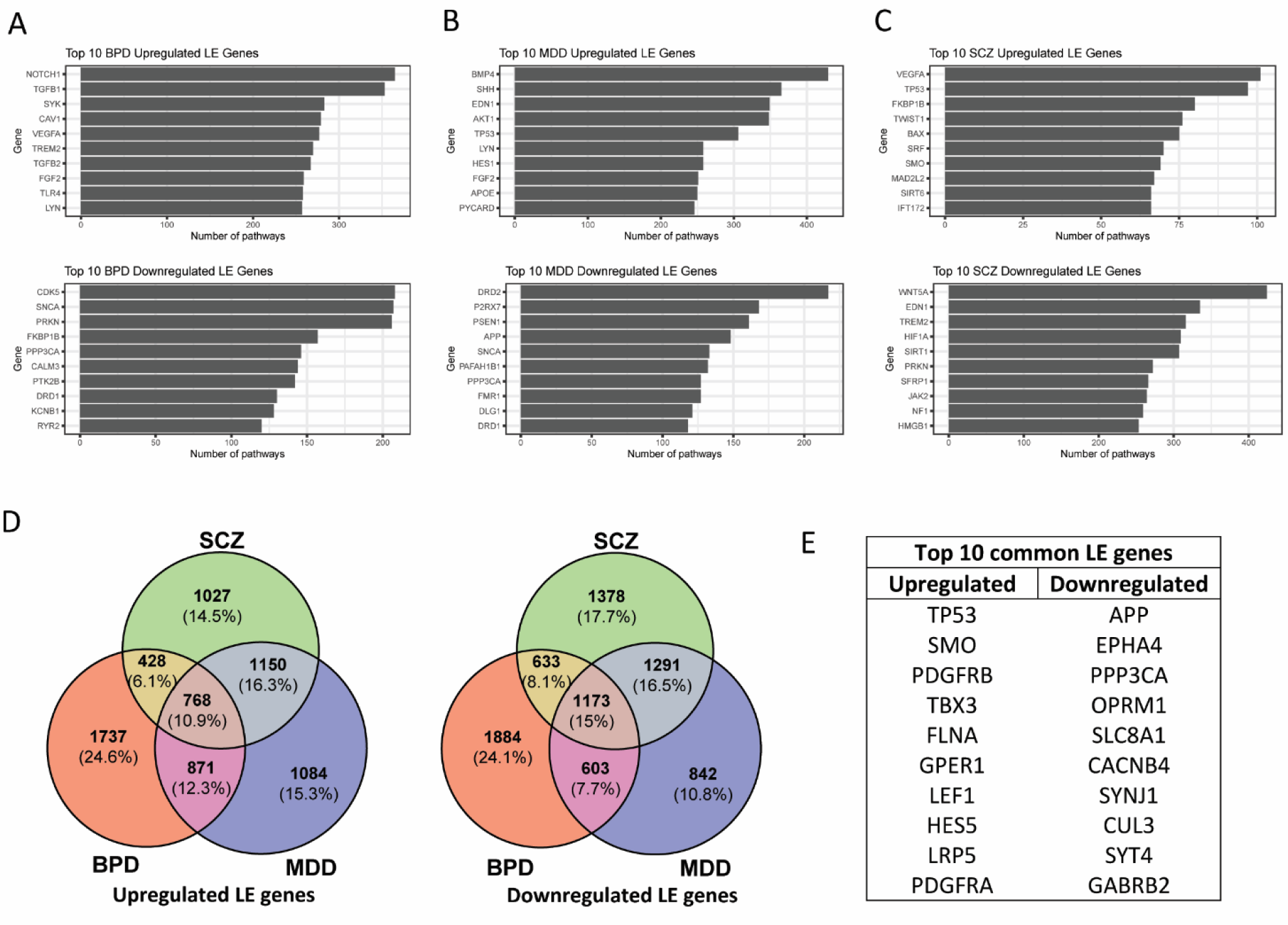
Leading edge gene analysis. The top 10 upregulated and downregulated leading edge (LE) genes, based on frequency identified in biological pathways are shown for BPD (**A**), MDD (**B**) and SCZ (**C**). (**D**) The number and proportion (%) of all LE genes and their intersection in BPD, MDD and SCZ. (**E**) The top 10 upregulated and downregulated LE genes that are common across BPD, MDD, SCZ. BPD bipolar disorder, MDD major depressive disorder, SCZ schizophrenia.

### LINCS chemical perturbagen analysis

iLINCS analysis identified chemical perturbagens (**Table S4**), organized by mechanism of action (MOA), that were dissimilar (discordant) or similar (concordant) to the transcriptional signatures of BPD (**Figure 4A-B**), MDD (**Figure S3A, C**) and SCZ (**Figure S3B, D**). The signature reversion principle suggests that chemical perturbagens that are discordant with the disease signature may induce gene expression changes that “reverse” disease-associated gene expression signatures ^55^. Equally, concordant chemical perturbagen signatures may indicate drugs that induce gene expression changes similar to those found in the disease state, informing on the underlying gene targets that may be implicated in disease.

**Figure 4.**
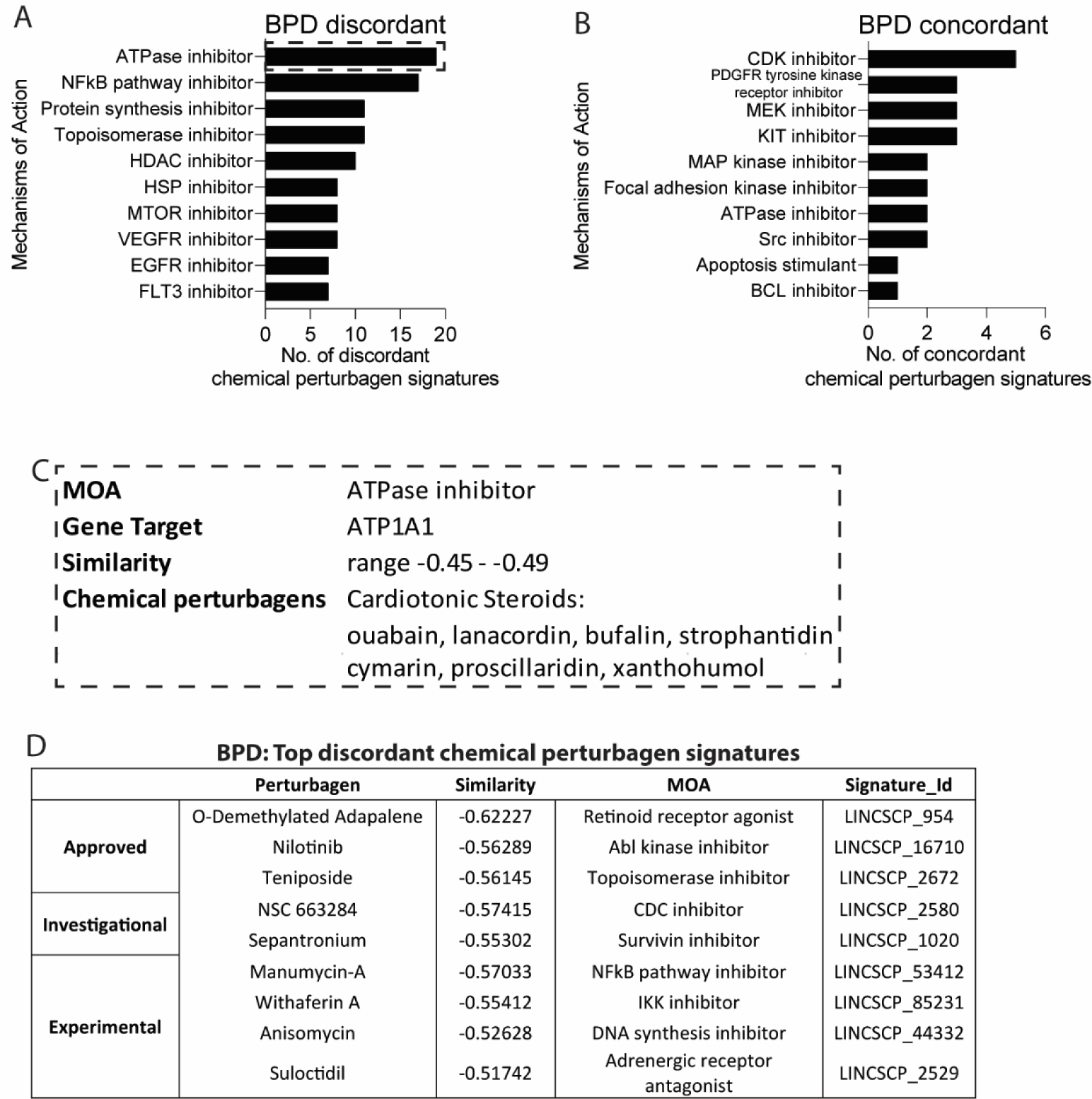
LINCS analysis. The mechanisms of action (MOA) of chemical perturbagens that are (**A**) discordant (dissimilar) and (**B**) concordant (similar) to the BPD amygdala LINCS signature. (**D**) The top BPD discordant MOA, ATPase inhibitor and the associated chemical perturbagens, cardiotonic steroids. (**E**) The top individual LINCS chemical perturbagen signatures that are most discordant with the BPD signature based on similarity score. BPD bipolar disorder, MOA mechanism of action.

Few chemical perturbagen signatures clustered by MOA were identified for SCZ and MDD analyses (**Figure S3**). However, “ATPase inhibitor” is the top mechanism of action for chemical perturbagens that are discordant to the BPD gene signature. The chemical perturbagens that comprise the nineteen ATPase inhibitor signatures include several cardiotonic steroids (**Figure 4D**) that modulate the Na^+^/K^+^ ATPase (ATP1A1-3) in a dose-dependent manner ^56^.

A small number of “ATPase inhibitor” chemical perturbagens (Blebbistatin (non-muscle myosin II ATPase inhibitor) and Evodiamine (ABCG2 inhibitor)) were identified as concordant with BPD, however these drugs do not target the Na^+^/K^+^ ATPase. ATP1A1 and ATP1A2 were significantly differentially expressed (p<0.05) in BPD but not SCZ and MDD. ATP1A1 was identified as a leading-edge gene in all three disorders, while ATP1A2 was identified as a leading-edge gene in BPD and MDD. We focused on the chemical perturbagens that share a common mechanism of action as this likely represents a more robust biological signature than single chemical perturbagen signatures (**Figure 4E**).

### Cell-type analysis

Cellular deconvolution analysis (**Figure 5**) identified significant reductions in cell type proportions of excitatory neurons in SCZ (p=0.004), BPD (p= 0.011) and MDD (p=0.013) relative to controls. Cell type proportions of astrocytes were significantly increased in SCZ (p=0.001), BPD (p=0.009) and MDD (p<0.001) compared to controls. Cell type proportions of oligodendrocytes were significantly reduced in SCZ (p=0.001) and MDD (p=0.019) but not BPD (p=0.2) compared to controls. These data support significant differences in cell compositions in psychiatric disorders compared to control.

**Figure 5.**
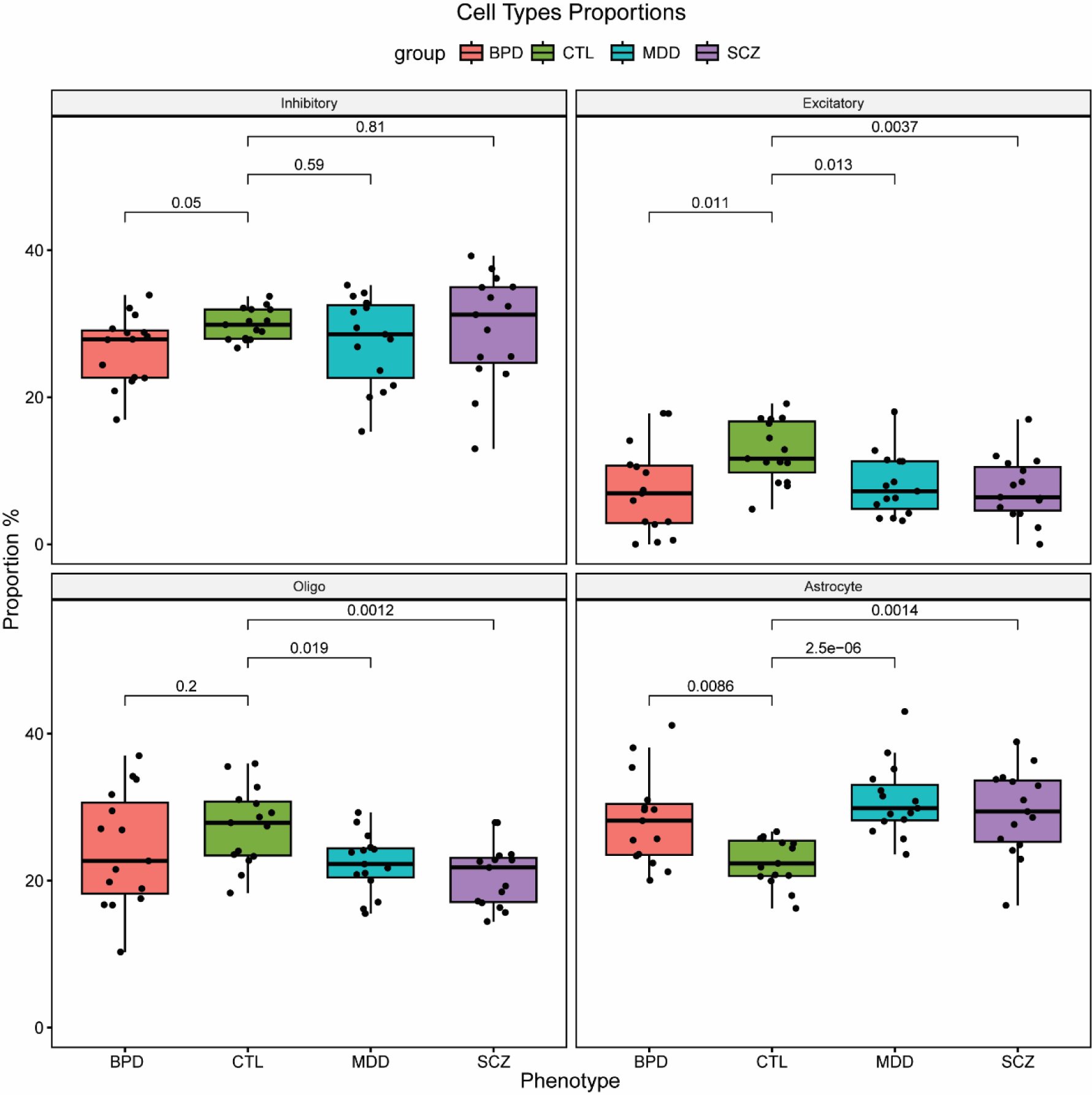
Cellular deconvolution. Cellular deconvolution from bulk RNAseq analysis of four different cell types in postmortem amygdala BPD, MDD and SCZ tissues. Cell type proportions (%) are shown for inhibitory and excitatory neurons, oligodendrocytes and astrocytes for BPD, MDD and SCZ relative to controls. BPD bipolar disorder, MDD major depressive disorder, MOA mechanism of action, SCZ schizophrenia.

### Medication effects

Antipsychotic medication was assessed as a potential confounding variable in variance partitioning analysis (**Figure S2**) however, it did not contribute significantly to the variance explained in the SCZ and BPD comparisons. We also compared the similarity of gene expression from our neuropsychiatric disorder RNAseq analysis with transcriptomic datasets obtained from the Stanley Online Genomics resource (https://www.stanleygenomics.org/) for SCZ subjects who were on and off antipsychotic medication and BPD subjects on and off mood stabilizers at time of death. Hypergeometric overlap analysis did not find significant overlap of the postmortem on/off medication and the psychiatric disorder datasets (p>0.05, **Figure S5**). An alternative statistic derived from hypergeometric overlap analysis, the Jaccard Index, which assesses the similarity between two sets of data, found that similarity was low (**Figure S5**, similarity 2% - 8%) suggesting the psychotropic medications do not drive the gene expression changes associated with disease. No dataset was available to assess the effects of medication on the transcriptome in MDD subjects who were on compared to off antidepressants. The medication datasets were not generated in amygdala tissue and thus may not reflect region-specific medication induced changes in gene expression.

## DISCUSSION

Psychiatric disorders share genetic and molecular pathologies ^57^ but the mechanisms that are common and unique to each disorder are still being elucidated. We present, to our knowledge, the first evidence for diagnosis-specific alterations of gene expression in metabolic pathways in the amygdala of subjects diagnosed with SCZ, BPD, or MDD. Furthermore, we identified biological pathways, including cell-cell signaling, biological adhesion, cell death, and calcium ion transport-related pathways that have unique patterns of enrichment in the amygdala in these disorders.

Our results indicate that metabolic pathway dysregulation in the amygdala is common across psychiatric conditions, but with distinct differences in specific metabolic signaling pathways in these disorders. Pathways involved in energy metabolism, for example “electron transport chain” and “oxidative phosphorylation” are downregulated in SCZ, upregulated in MDD but not significantly enriched in BPD. In addition, nucleotide metabolism pathways like “purine nucleotide metabolic process” are significantly downregulated in SCZ. Deficits in mitochondrial bioenergetics are widely reported in the brain in SCZ ^58^. Reduced transcript and protein expression of electron transport chain complex I and III enzymes ^59^, and lower activity of complex IV ^60^ were found in the frontal cortex in subjects diagnosed with SCZ. Similar reductions in mitochondrial complex activity were found in the temporal cortex and basal ganglia ^60^. Decreases in the transcript expression of mitochondrial genes and associated biological pathways were also reported in the hippocampus ^61^, in superficial and deep lamina of the dorsolateral prefrontal cortex (DLPFC) ^62, 63^, and in enriched populations of laser captured parvalbumin interneurons in the DLPFC ^64^. Our findings in the amygdala lend further support for downregulation of bioenergetic processes across different brain regions in SCZ. In contrast, we found bioenergetic pathways were upregulated in the amygdala in MDD subjects. Increased levels of mitochondrial transcript expression ^65^ and increased protein expression of twenty different subunits of the oxidative phosphorylation complex were reported in the DLPFC in MDD subjects ^66^. However, as ATP levels were found to be reduced in the DLPFC in these subjects, upregulation of oxidative phosphorylation complexes may represent a compensatory response to overcome energy deficits in MDD ^66^. Although significant changes in mitochondrial transcript expression in postmortem MDD brain tissue were found after controlling for the effects of antidepressant treatment ^65^, in rodent models, administration of selective serotonin reuptake inhibitors (SSRIs) resulted in the upregulation of proteins implicated in energy metabolism and ATP synthesis ^67^. However, positron emission tomography (PET) imaging with [(18)F] fluorodeoxyglucose found elevated anterior cingulate metabolism levels and reduced prefrontal metabolic activity that were normalized following administration of the antidepressant paroxetine in MDD patients ^68^. This suggests basal, brain-region specific dysregulation of energy metabolism in MDD that is amenable to pharmacotherapy.

Conversely, carbohydrate metabolism-related pathways, including “positive regulation of carbohydrate metabolic process” and “monocarboxylic acid metabolism” are upregulated in BPD, with few pathways enriched in SCZ and MDD in the amygdala. PET studies using 8F-fluorodeoxyglucose have previously identified increased glucose metabolism in the left amygdala in bipolar-depressed patients ^30, 69^. Emerging evidence suggests that circumventing glycolysis may be an effective therapy for psychiatric disorders, particularly BPD. Ketogenic diets replace brain carbohydrate metabolism with ketones as an energy source ^70, 71^ and show promise in treating symptoms in people with BPD and SCZ ^20, 25, 72–74^. Altered glycolysis was also reported in the amygdala in depression patients ^75^, but we did not identify significant enrichment of these energy metabolism pathways at the transcriptomic level in this study. Widespread bioenergetic dysregulation, including glucose utilization deficits ^58, 76, 77^, availability of ATP reserves ^78–81^ and mitochondrial enzyme dysfunction ^82, 83^ have been identified in postmortem brain tissue studies and imaging studies of psychiatric patients, although fewer studies have been conducted in the amygdala. Our results also support metabolic dysfunction as a core feature of psychiatric disorders, but with disease and brain-region specific perturbations in bioenergetic processes.

Interestingly, the primary finding from iLINCS chemical perturbagen analysis is that drugs that regulate the Na^+^/K^+^ ATPase (ATPase inhibitors) are discordant with the BPD disease signature. The ATPase inhibitors identified here were predominantly cardiotonic steroids, which modulate the activity of the Na^+^/K^+^ ATPase in a dose-dependent manner ^84^. Na^+^/K^+^ ATPase maintains plasmalemma membrane potential in neurons by reestablishing Na^+^ and K^+^ ion gradients following action potential firing. The Na^+^/K^+^ ATPase enzyme is posited to be the single largest consumer of ATP in the brain ^85^, and is an important regulator of ion homeostasis ^86^. Preclinical models targeting the Na^+^/K^+^ ATPase indicate that inhibition of this enzyme may contribute to manic symptoms in BPD. For example, intracerebroventricular administration of the ATPase inhibitor ouabain reduces Na^+^/K^+^ ATPase pump activity and increases dopamine release and locomotor activity in rats, which are alleviated by lithium administration ^87, 88^. ILINCS analysis identified ATPase inhibitors as compounds that may reverse amygdala BPD transcriptional signatures. This finding seems contradictory, as Na^+^/K^+^ ATPase inhibition is associated with BPD symptoms. However, endogenous cardiotonic steroids can have different physiological effects ^89^, and at low doses, cardiotonic steroids can increase activity of the Na^+^/K^+^ ATPase ^90, 91^. We found that expression of the primarily neuronally-expressed Na^+^/K^+^ ATPase subunit gene ATP1A1 ^92^ was downregulated in BPD but expression of the glial-cell expressing subunit ATP1A2 was significantly upregulated. Variable levels of expression of different Na^+^/K^+^ ATPase α isoforms have previously been reported in the prefrontal cortex ^93^, temporal cortex ^94, 95^ and parietal cortex ^96^ in BPD. The complex role of Na^+^/K^+^ ATPase, the functional effects of changes in expression of its different isoforms and its potential role as a disease mechanism in BPD are discussed in detail elsewhere ^97–99^. However, our results lend support to the hypothesis that modulation of Na^+^/K^+^ ATPase activity may play a role BPD pathophysiology and serve as a therapeutic target ^100^.

We observed selective downregulation of calcium ion signaling in subjects with BPD, along with several genes involved in L-type calcium channel signaling as leading-edge genes including CACNA1C and CACNB2 (**Figure 2**). CACNA1C polymorphisms are one of the most strongly implicated genetic factors in GWAS studies of BPD ^101–107^ and represent a promising factor for developing personalized treatments. A recent GWAS also identified enrichment of polygenic risk factors for targets of calcium channel blockers, including the L-type calcium channel blocker isradipine ^108^. Previous studies have reported increased fMRI activity as well as CACNA1C mRNA expression in the dorsolateral prefrontal cortex of control subjects with two copies of the risk allele ^109^. Furthermore, increased mRNA expression along with L-type calcium current was reported in cultured induced neurons from people with two copies of the CACNA1C risk allele ^110^. In contrast, two studies have reported decreased CACNA1C mRNA expression in the superior temporal gyrus and the cerebellum in people with the risk allele ^111, 112^. These discrepancies in gene expression levels may be due to variability in multiple CACNA1C polymorphisms between cohorts, brain region specific effects including somatic mutations or medication effects.

Despite the evidence from genetic association studies ^101–108^, the iLINCS perturbagen analysis did not identify calcium channel blockers as top discordant drugs for the gene expression signatures in any diagnosis group. However, ATPase inhibitors such as the cardiotonic steroid ouabain were the top discordant drugs identified for subjects with BPD. In addition to bioenergetic perturbations, enrichment of chemical perturbagens that modulate ion pumps like the Na^+^/K^+^ ATPase in BPD ^113^ lends further support for ion dysregulation as a common pathological feature of this disorder ^114^. ATPase inhibitors regulate cellular Na^+^ and K^+^ ion levels via their action at the Na^+^/K^+^ ATPase pump but can also increase intracellular calcium levels and in turn activation of several cell signaling pathways ^115^. Increased intracellular calcium has been reported in subjects with BPD ^116–118^. Furthermore, ATPase inhibitors can act as anti-inflammatory compounds ^119, 120^. Immune system processes were selectively upregulated in the amygdala of subjects with BPD, which may also have contributed to the identification of ATPase inhibitors as discordant with the BPD disease-related gene signatures. The subjects diagnosed with BPD in our study likely represent subjects in a depressed or euthymic state, suggested by the prevalence of suicide in this group. The decreased L-type calcium signaling we observed in our study may be associated with depressive states whereas increased L-type calcium signaling may reflect manic episodes. Calcium channel blockers have been reported to be effective in treating mania and to a lesser extent depression in patients with BPD ^121–126^. Collectively, altered ATPase and L-type calcium channel pathways may be at the center of energy balance dysfunction in BPD, reflected by increased activity during mania and decreased activity in depression.

Biological adhesion pathways, including CD4 positive T-cell pathways, were enriched only in the amygdala of subjects with SCZ. Pathways involved in immune system processes were downregulated in subjects with SCZ. These findings contrast with reports of increased cell adhesion and immune system molecules in serum and cortical brain samples from subjects with SCZ ^127, 128^. Cell adhesion alterations and immune system dysregulation are associated with metabolic perturbations in SCZ. Cell adhesion molecules are differentially expressed in serum from SCZ patients diagnosed with comorbid metabolic syndrome compared to SCZ patients without metabolic syndrome ^129^. Metabolic differences in each cohort, as well as brain region specific changes in cell adhesion processes, may account for the differences in alterations in these pathways in our findings compared with previous reports.

Our previous studies identified alterations in extracellular matrix molecules (ECMs) and perineuronal nets in the amygdala of subjects with SCZ, with more moderate changes in BPD ^35, 130^. In the current study ECM pathways including “diseases associated with glycosaminoglycan metabolism”, “proteoglycans in cancer”, “vasculature development” and “blood vessel morphogenesis” were enriched in subjects with BPD, compared to a lack of enrichment of ECM pathways in SCZ or MDD. This included the ECM genes PTPRZ1, VCAN, ST6GAL1, SEMA3G and NCAN in subjects with BPD. GWAS studies reported a genetic polymorphism in NCAN associated with BPD ^131^, and human and mouse studies suggest this genetic factor is involved in manic symptoms ^132^. We identified a small number of differentially expressed ECM genes in subjects with SCZ including ST6GALNAC4, COL1A1, and COL1A2. Our results may reflect more subtle alterations in ECM pathways in the amygdala of subjects with BPD that may contribute to the previously reported alterations of PNN composition in this region ^130^.

Transcriptional pathways involved in “cell death”’ were markedly and selectively upregulated in subjects with MDD and consisted of apoptotic and programmed cell death pathways. Apoptosis-related gene transcript ^133^ and protein marker expression ^134^ are found in the frontal cortex in MDD, suggesting increased vulnerability to persistent low-grade cell degeneration in this disorder. In the amygdala, volume changes are associated with reduced glial cell numbers, particularly oligodendrocytes ^135^. In line with this, we also found reduced cell proportions of oligodendrocytes in MDD subjects, along with reduced excitatory neuron proportions and increased proportions of astrocytes. Upregulated cell death pathways in MDD may preferentially impact these two cell types. As in MDD, increased cell-type proportion of astrocytes was also observed in BPD and SCZ, supporting a role for glial cell dysfunction in these disorders that may contribute to disease-specific transcriptomic changes ^136^. For example, increased astrocyte proportions may contribute to inflammation, as suggested by upregulated immune system processes in BPD. In comparison, increased astrocyte cell proportions in SCZ together with downregulated immune system processes suggest impaired astrocytic function in this disorder. Fewer immune related pathways were identified in MDD compared to the other psychiatric disorders. Immune system dysregulation plays an important role in MDD ^137^ but our findings suggest it is not a major driver of pathological changes in the amygdala.

A limitation of cellular deconvolution is that analysis is carried out with the assumption that up and downregulated cell-type specific marker expression is coordinated and reflects proportional increases or decreases of a cell type ^138^. That is, analysis does not distinguish between increased cell number and higher expression of cell markers for that cell type. It also does not consider the cellular reactivity state which may be relevant for the astrocyte cell subtype. Future studies using single nucleus RNAseq or high-resolution microscopy mRNA and protein analysis will provide more insight into the cell-type specific alterations in the transcriptomic pathways identified in our study. The information available for our cohort did not allow for analysis of potential relationships of mania, depression, and euthymia at death on our outcome measures. Future studies using larger cohorts with more detailed patient histories may shed light on the specific effects that mood state may have on metabolic pathways and ATPase signaling.

In summary, our results identify diagnosis-specific alterations in metabolic pathways, immune pathways, and calcium ion transport in the amygdala of subjects with SCZ, BPD, and MDD. These results suggest that the amygdala is a region where alterations in these pathways occur in a disease-specific manner, potentially associated with broader metabolic and immune system dysfunction that is often comorbid with these disorders. Identifying disease-specific alterations may guide the development and application of metabolic-based therapeutic strategies for SCZ, BPD, and MDD.

## Acknowledgements

Postmortem brain tissue was donated by The Stanley Medical Research Institute Brain Collection. We would like to thank Maree J. Webster for her assistance with use of this sample collection for our study.

## Data Availability Statement

The data and original contributions presented in this study are included in the manuscript or supplemental materials. All gene expression data is available from the <u>Stanley Neuropathology Consortium Integrative Database</u>. Further inquiries can be directed to the corresponding authors.

