## Supplemental Figures for "Differentially Altered Metabolic Pathways in the Amygdala of Subjects with Schizophrenia, Bipolar Disorder and Major Depressive Disorder"

**Supplementary Figures**

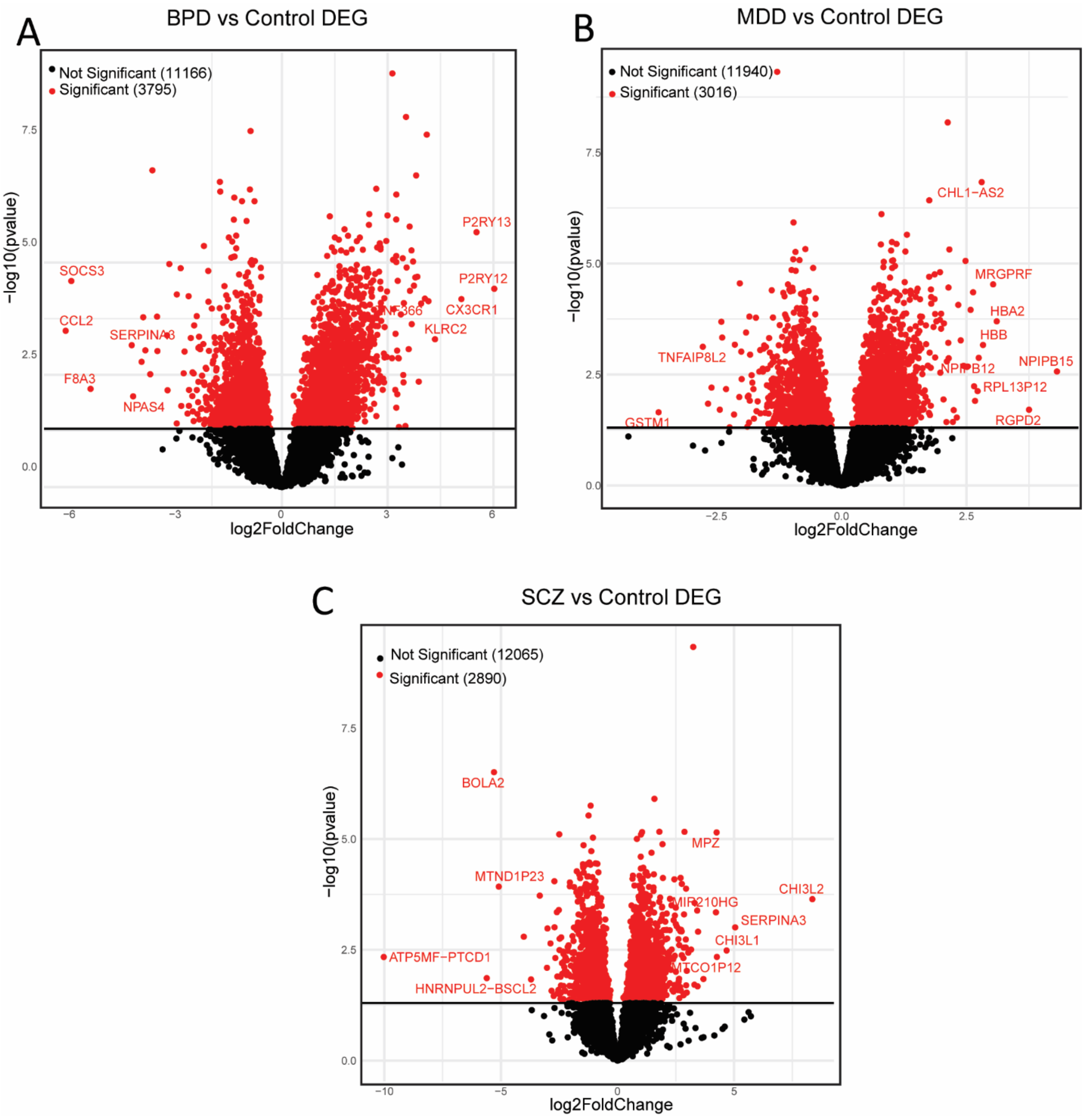


**Fig S1. Volcano Plot.** Volcano plot of -log10(p-value) (y-axis) and log2 fold change (x-axis) of differentially expressed gene transcripts in BPD (**A**), MDD (**B**) and SCZ (**C**). Significant genes are indicated by red dots, above the horizontal line indicating significance threshold (-log10(p<0.05)). Top differentially expressed genes (DEGs) are named on each figure. BPD bipolar disorder, MDD major depressive disorder, SCZ schizophrenia.

**Figure S2**

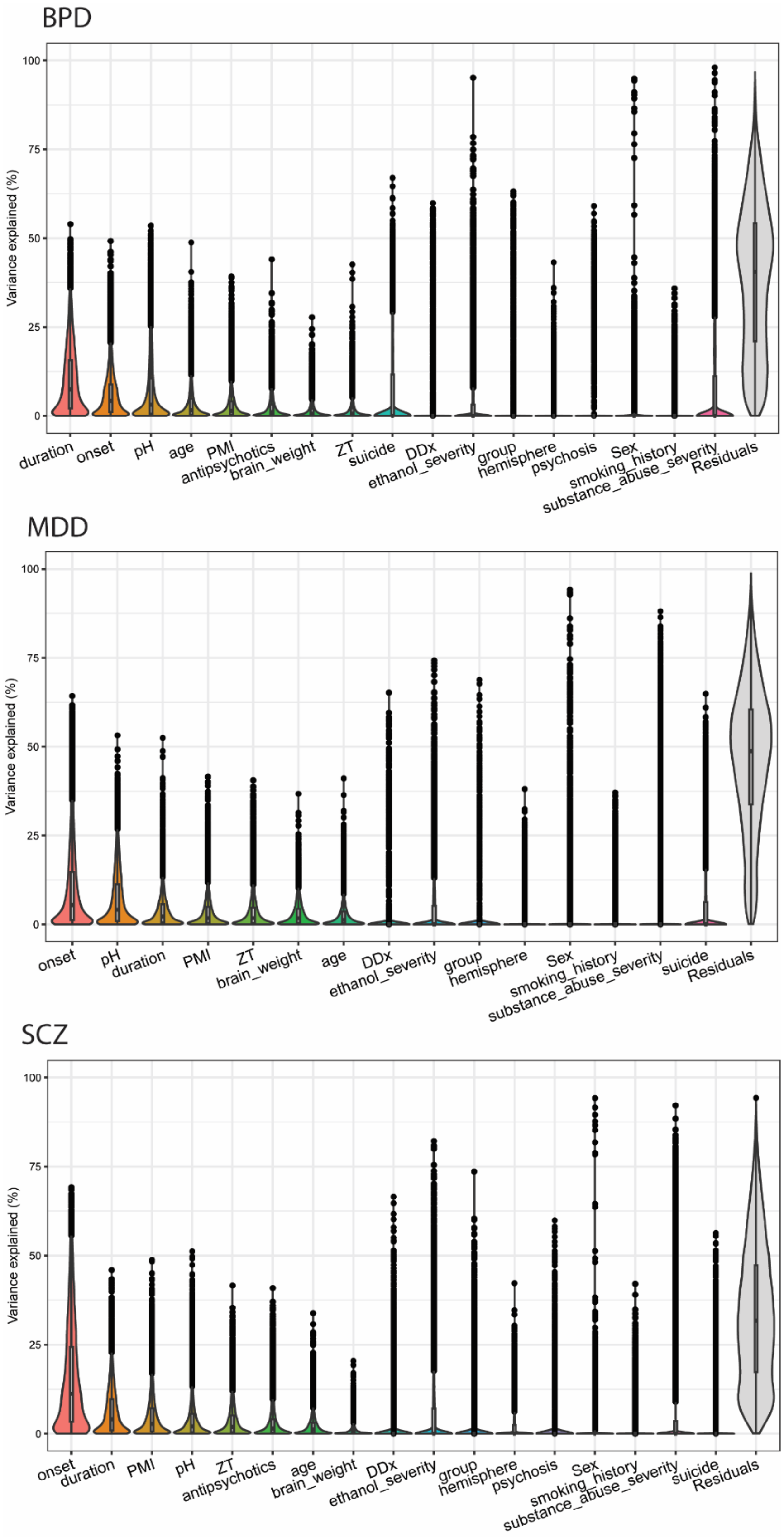
BPD
Variance explained (%)
100
75
50
25
0
duration
onset
pH
age
PMI
antipsychotics
brain_weight
ZT
suicide
DDx
ethanol_severity
group
hemisphere
psychosis
Sex
smoking_history
substance_abuse_severity
Residuals
MDD
Variance explained (%)
100
75
50
25
0
onset
pH
duration
PMI
ZT
brain_weight
age
DDx
ethanol_severity
group
hemisphere
Sex
smoking_history
substance_abuse_severity
suicide
Residuals
SCZ
Variance explained (%)
100
75
50
25
0
onset
duration
PMI
pH
ZT
antipsychotics
age
brain_weight
DDx
ethanol_severity
group
hemisphere
psychosis
Sex
smoking_history
substance_abuse_severity
suicide
Residuals

**Fig S2. Variance partitioning analysis.** Violin plots of all covariates (x-axis), ranked by percentage (%) variance explained (y-axis) for BPD, MDD and SCZ. The top 2 covariates were included as covariates in generalized linear regression model analyses for all disease comparisons. BPD bipolar disorder, DDx diagnosis, MDD major depressive disorder, PMI postmortem interval, SCZ schizophrenia, ZT zeitgeber time.

**Figure S3**

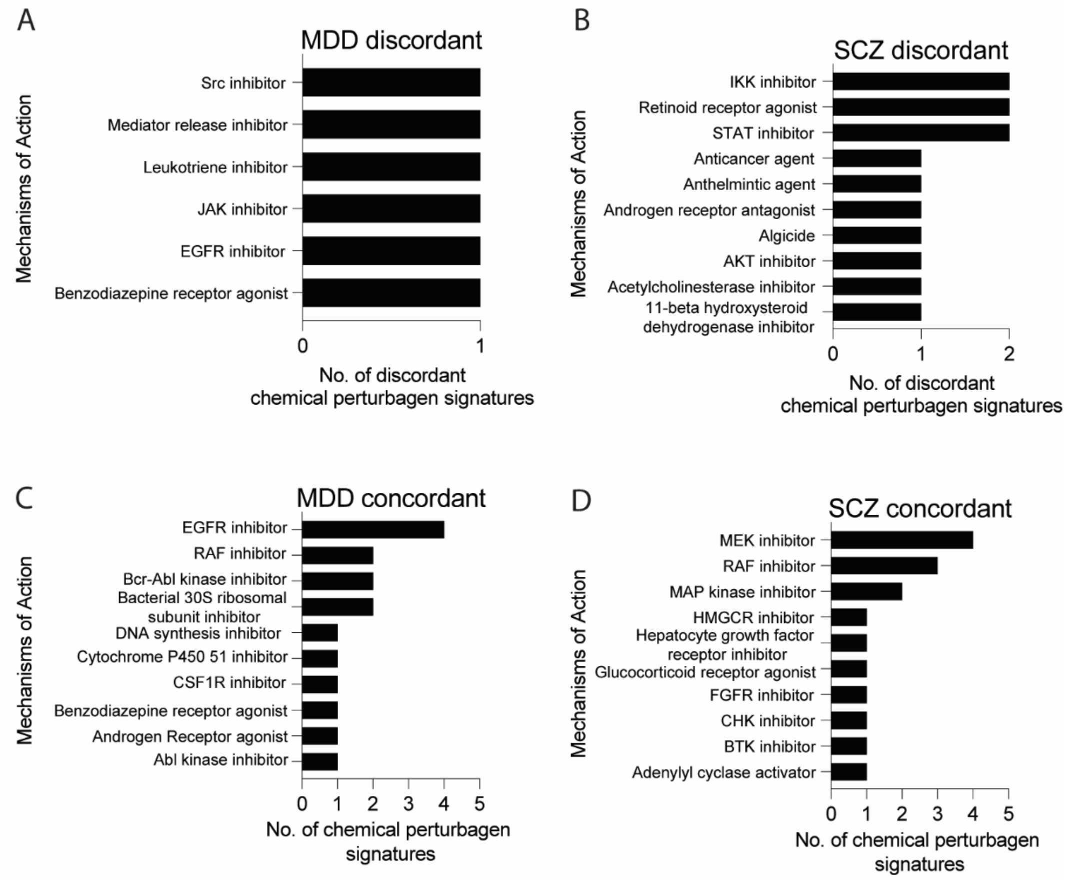


**Fig S3. LINCS analysis MDD and SCZ.** The mechanisms of action (MOA) of the chemical perturbagens with LINCS signatures that are discordant (dissimilar) to the MDD **(A)** and SCZ **(B)** disease signatures or concordant (similar) to the MDD **(C)** and SCZ **(D)** disease signatures. MDD major depressive disorder, MOA mechanism of action, SCZ schizophrenia.

**Figure S4**

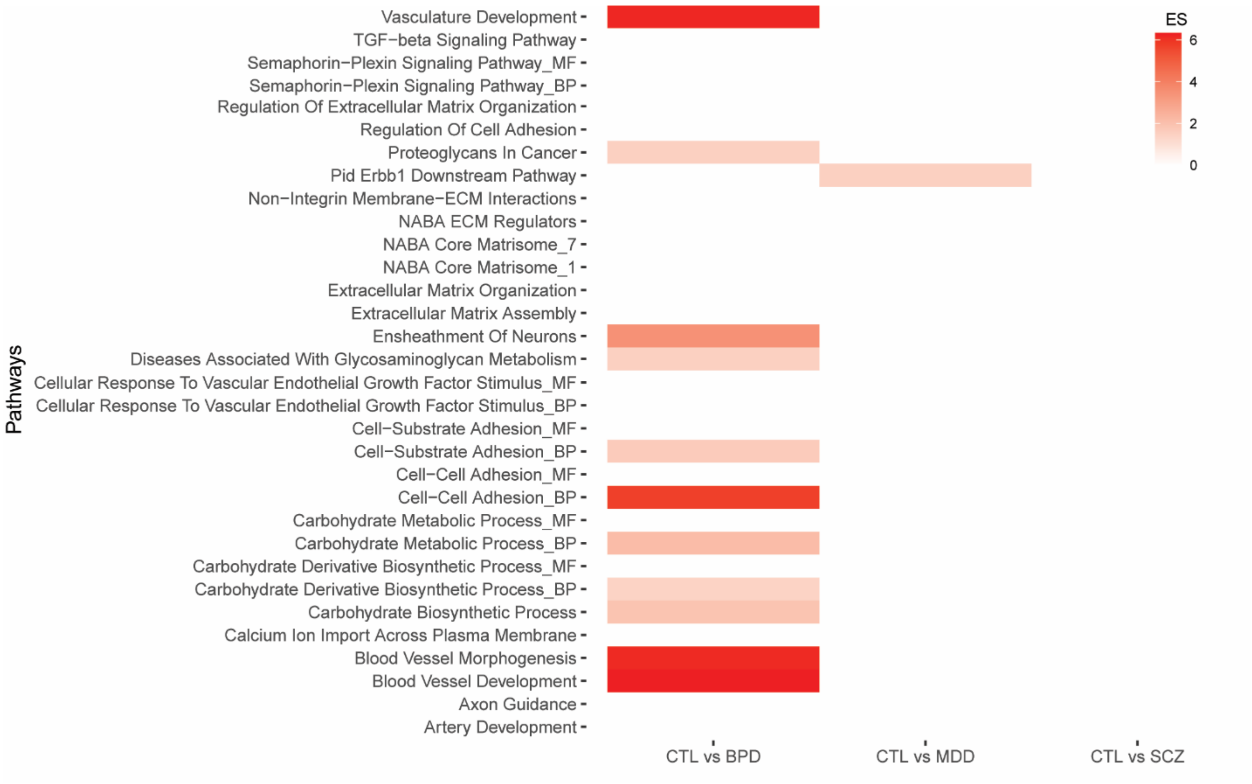


**Fig S4. Heatmap overlap**. Hypergeometric overlap analysis of metabolic genesets and amygdala transcriptional profiles from BPD, MDD and SCZ.

**Figure S5**

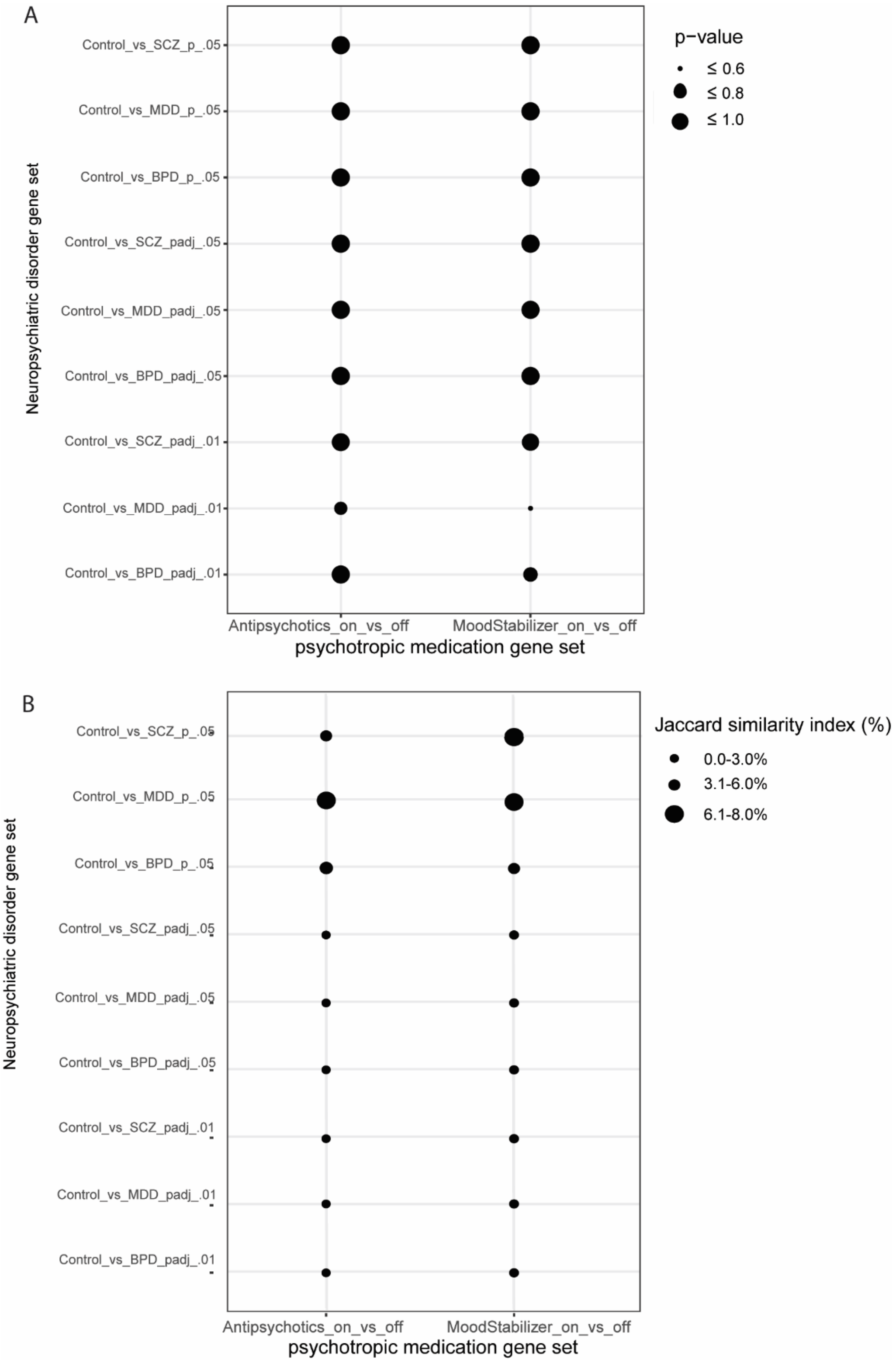


**Fig S5. Psychotropic medication gene set enrichment. (A)** Hypergeometric overlap analysis found no significant overlap ($p > 0.05$ all comparisons) of BPD, MDD and SCZ DEGs (psychiatric disorder gene sets composed of DEGs at FDR<0.01, FDR<0.05, and p-value <0.05) and DEGs ($p < 0.05$) from subjects who were on v off antipsychotics or on v off mood stabilizers at time of death (obtained from the Stanley Medical Research Institute Online Genomics Database). **(B)** The Jaccard similarity index, which calculates the similarity of two gene sets ranging from 0 - 100%, was also generated for the same datasets. No significant overlap of DEGs altered in on/off medication datasets were found using either approach.
